# Modified Oxacillin Resistant *Staphylococcus aureus* detected in neonatal intensive care patients

**DOI:** 10.1101/2021.03.04.21251785

**Authors:** Melissa R. Gitman, Bremy Alburquerque, Adriana van de Guchte, Mitchell J. Sullivan, Ajay Obla, Jose Polanco, Marilyn Chung, Irina Oussenko, Melissa L. Smith, Flora Samaroo, Debbie Barackman, Deena R. Altman, Emilia Mia Sordillo, Harm van Bakel

## Abstract

Active surveillance in our neonatal intensive care unit identified *Staphylococcus aureus* cultures from two infants with heterogeneity in methicillin resistance between isolated subclones lacking *mecA* and *mecC*. Whole-genome analysis of 4 modified (MODSA) and 4 methicillin-susceptible (MSSA) subclones for each culture identified either truncating mutations in the cyclic diadenosine monophosphate phosphodiesterase enzyme (GdpP), or a nonsynonymous substitution in penicillin binding protein 2 (PBP2). These cases highlight the difficulty in identifying non-*mecA/*non-*mecC*-mediated methicillin-resistance in clinical laboratories.

## Background

Expression of MecA or MecC is the most commonly recognized mechanism for methicillin-resistant Staphylococcus aureus (MRSA) [1]. Non-MecA and non-MecC mediated resistance through other mechanisms also has been described [1,2]. When these involve isolates with mutations to native genes such as those encoding penicillin binding proteins (PBPs) [3–7], GdpP [1,2,6,8,9], the cation multidrug efflux transporter AcrB [6] or the thioredoxin-related protein YjbH [10], they are generally referred to as modified *S. aureus* (MODSA). Alternatively, resistance caused by overproduction of β-lactamases is typically classified as borderline resistant *S. aureus* (BORSA) [1]. Methods of detection for MODSA or BORSA are limited. Here we describe the use of whole-genome sequencing to determine the mechanisms of methicillin resistance for two patients with heterogeneous MSSA/MODSA surveillance cultures, highlighting the challenges of identifying these organisms in a routine clinical microbiology laboratory.

## Methods

### Ethics statement

The study protocol was reviewed and approved by the Mount Sinai School of Medicine Institutional Review Board for the collection and bacterial genome sequencing of discarded clinical specimens by the Pathogen Surveillance Program (protocol HS# 13-00981), as defined by DHHS regulations. A waiver of authorization for use and disclosure of protected health information (PHI) and a waiver of informed consent was approved based on the criteria that the use or disclosure of PHI involved no more than minimal risk to the privacy of individuals, and because the research could not practically be conducted without the waiver and without access to and use of the PHI. The research conformed to the principles of the Helsinki Declaration.

### Phenotypic testing

Isolates were processed per the Clinical Microbiology Laboratory (CML) Standard Operating Procedure for screening for MRSA. Swabs were plated on chromID® MRSA agar (CA) (bioMérieux, Marcy-l’ Etoile, France) and on trypticase soy agar supplemented with 5% sheep blood (SBA) (BD Diagnostics, Sparks, MD) and incubated at 37°C for 24 hours. Beta-hemolytic colonies on SBA were identified via conventional biochemical testing. Isolates that were catalase- and coagulase-positive were confirmed and tested for antimicrobial susceptibility by automated broth microdilution assays on the Vitek 2 platform (bioMérieux, Marcy-l’Etoile, France), as well as cefoxitin disk screening (cefoxitin 30 μg; BD Sensi-disc, Becton Dickinson, Germany).

Additional testing of MSSA/MODSA subcultures was done using the latex agglutination Staphaurex™(Thermo Fisher Scientific, Switzerland), mannitol salt agar (BD Diagnostics, Sparks, MD), and the Cepheid Xpert® SA Nasal Complete (Cepheid, Sunnyvale, CA, USA). Phenotypic testing of subcultures was performed using the Vitek® GP 2 ID panel, the Gram Positive Susceptibility PM34 panel on the MicroScan platform (Beckman Coulter, CA, USA), as well as cefoxitin disk, and oxacillin E-test (bioMérieux SA, Marcy L’Etoile, France) using Mueller-Hinton agar supplemented with 2% NaCl (BD Diagnostics, Sparks, MD) as per Clinical and Laboratory Standards Institute (CLSI) performance standards. (11)

### DNA preparation and sequencing

For genome sequencing, selected subclones were cultured on SBA (Thermo Fisher Scientific) under nonselective conditions. The Qiagen DNeasy Blood & Tissue Kit (Qiagen, Hilden, Germany) was used for DNA extraction, as previously described [12]. Following quality control, DNA and library preparation, long-read sequencing was performed on the Pacific Biosciences RS-II platform to a depth of >200x. Additional Illumina short-read sequencing was performed to aid final assembly polishing. Briefly, libraries were prepared for DNA isolated from the same subclones using the Nextera DNA Flex Library Prep Kit (Illumina), and sequenced in a paired-end format (2×150 nt) on the NextSeq Platform to a minimum depth of 86 fold.

### Genome assembly and comparison

PacBio long-read sequencing data were processed using a custom genome assembly pipeline, as previously described [12]. Complete and circularized genome assemblies were then polished using the arrow algorithm [13] based on minimap2-aligned [14] PacBio raw subreads. Next, Illumina paired-end reads were aligned to PacBio assemblies using BWA-MEM [15] followed by a second round of assembly polishing with Pilon [16]. Finally, high-quality curated genomes were annotated using PROKKA [17] and visualized using the Integrated Genome Browser [18]. Pairwise-comparisons between the genomes of susceptible and resistant samples were performed using NucDiff [19] to identify genomic variants that correlated with sample phenotypes, and the functional effect of these genomic variants was annotated using ANNOVAR [20]. Genic regions annotated as encoding hypothetical proteins by PROKKA were subjected to additional BLASTP searches [21] to attempt to identify these proteins.

## Results

During NICU surveillance for MRSA colonization of the nares, isolates from two infants (i1 and i2) yielded green colonies on CA plates. Automated broth microdilution testing confirmed both isolates to be MRSA. However, when the same isolates were subcultured and sequenced for research purposes, both subcultures (named i1-SC0 and i2-SC0) were identified as MSSA on CA plates and by comparing the resulting genomes to earlier MSSA isolates obtained from the same infants.

To investigate the cause of the discrepancy between the clinical and research assays, we performed additional evaluation of the original surveillance cultures. Isolates from both infants were plated again on SBA and CA, and incubated at 37°C. After 24 hours, plates were reviewed and 2 colonies of different morphologies from the CA and 3 colonies from the SBA were selected, and subcultured to CA and SBA, for a total of 5 subcultures per infant. Each subculture, including the original subcultures obtained for research purposes, were then tested using Staphaurex, mannitol salt agar, and the Cepheid Xpert MRSA assay. All subcultures from both infants were confirmed to be *S. aureus*, and tested negative by the Cepheid Xpert MRSA assay for *mecA, mecC*, and SCC*mec*. Repeat phenotypic testing was performed for all 12 isolates using the Vitek® GP 2 panel or the PM34 MicroScan panel, as well as cefoxitin disk, and oxacillin E-test using Mueller-Hinton agar supplemented with 2% NaCl as per CLSI methods. For i1, 3 of 6 subcultures tested consistently as MRSA, and 3 as MSSA (**Table 1**). For i2, only one of 6 subcultures tested as MRSA based on cefoxitin screen and oxacillin E-test, although but not by automated oxacillin testing (**Table 1**).

**Table 1.** Phenotypic Antimicrobial Susceptibility Testing Results.

| Infant 1 (i1) |  |  |  |  |  |  |
| --- | --- | --- | --- | --- | --- | --- |
| Test | SC0 | SC1 | SC2 | SC3 | SC4 | SC5 |
| Cefoxitin (ABM) | NEG | POS | POS | NEG | NEG | POS |
| Cefoxitin disk | S | S | S | S | S | S |
| Oxacillin (ABM) | S (1) | R (>=4) | R (>4) | S (0.5) | S (0.5) | R (>4) |
| Oxacillin E-test | S (1) | R (8) | R (8) | S (0.5) | S (0.5) | R (8) |
| Infant 2 (i2) |  |  |  |  |  |  |
| Test | SC0 | SC1 | SC2 | SC3 | SC4 | SC5 |
| Cefoxitin (ABM) | NEG | NEG | NEG | POS | NEG | NEG |
| Cefoxitin disk | S | S | S | S | S | S |
| Oxacillin (ABM) | S (0.5) | S (<=0.25) | S (0.5) | R* (0.5) | S (<0.25) | S (0.5) |
| Oxacillin E-test | S (1.5) | S (0.25) | S (0.19) | R (8) | S (0.25) | S (0.25) |

ABM = automated broth microdilution assays on the Vitek or Microscan platform; SC = subclone; S = susceptible; R = resistant; R\* = converted via expert rule to resistant; POS = positive; NEG = negative. Minimal inhibitory concentration (MIC) is indicated between brackets where available.

The finding of susceptible and resistant subclones in the surveillance cultures from the same individuals suggested the presence of genetic heterogeneity. As such, we performed whole-genome sequencing and assembled finished-quality genomes for three resistant and two susceptible subclones for i1, as well as one resistant and two susceptible subclones of i2. Comparison of complete genomes identified 8 variants among i1 subclones and 10 variants among i2 subclones (**Table S1**). In i1, only the gene encoding *gdpP* was mutated in a pattern that was consistent with the observed phenotypic differences. Each of the three resistant isolates had a different *gdpP* mutation; namely a truncating frameshift deletion at M613fs, a substitution at G291V, or a truncating frameshift insertion at V371fs (**Figure 1A**). For i2, the resistant isolate was found to have a T552I substitution in the transpeptidase domain of Penicillin Binding Protein 2 (PBP2), as well as a A574T substitution in RpoB (**Figure 1B**).

**Figure 1.**
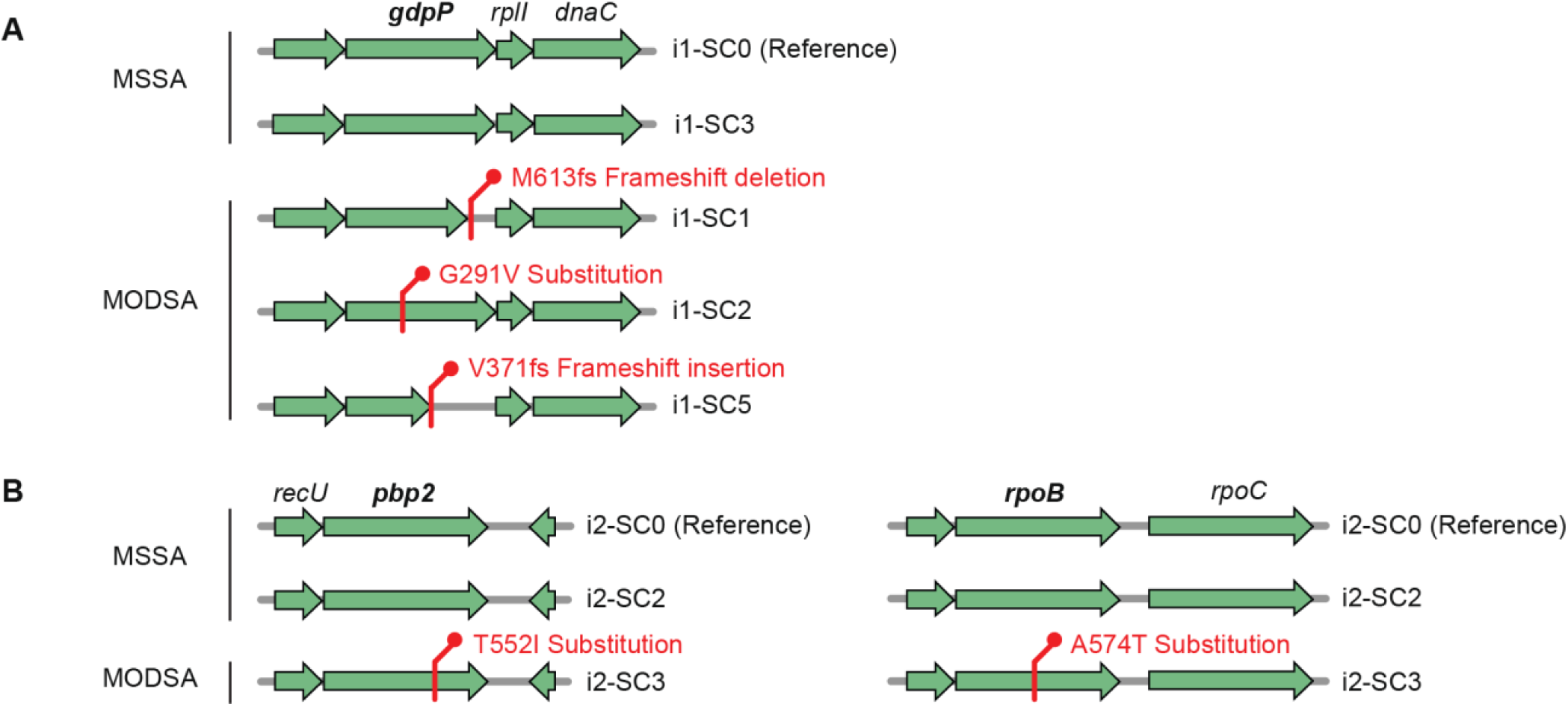
Genetic variants uniquely identified in MODSA strains. **A)** Variants seen in MODSA strains in Infant 1 that were not present in MSSA strains from the same patient. **B)** Same as A but for Infant 2.

## Discussion

In this study we identified 2 *S. aureus* isolates of a mixed MODSA MSSA phenotype that did not contain *mecA* or *mecC*. The resistant isolates from i1 contained variants in the gene encoding GdpP. It has been previously described that GdpP hydrolyses the secondary messenger cyclic diadenosine monophosphate (c-di-AMP) [8], such that inactivation or deletion of *gdpP* leads to an increased level of c-di-AMP, which produces a reduced susceptibility to β-lactams [2,8,9]. In a study by Ba et al., an array of truncating mutations of the *gdpP* gene was identified in 17 out of 20 *mec*-negative isolates [1]. Moreover, when a plasmid copy of the gene was reinserted in 4 isolates, susceptibility to oxacillin was restored [1]. What is unusual in our patient’s isolates is that although a diversity of mutations in the gdpP gene have been previously described, to our knowledge this is the first report of the concurrent isolation and identification from an individual patient of 3 different subclones with different *gdpP* mutations in the same strain. In i2, we found only one subclone that expressed phenotypic methicillin resistance, with mutations in *pbp2* and *rpoB. Staphylococcus aureus* possesses four penicillin-binding proteins (PBPs) that contribute to the assembly of cell wall peptidoglycan. However, functional PBP2 has an important role in the expression of resistance to methicillin [5,22], and the T552I substitution observed in the transpeptidase domain of PBP2 has previously been identified in *mec*-negative methicillin resistant *S. aureus* isolates [7]. There is also evidence to suggest that *rpoB* mutations may contribute to MRSA [23, 24]. Aiba et al. demonstrated that introduction of an *rpoB* mutation was accompanied by tolerance to bactericidal concentrations of methicillin [23]. Moreover, Panchal et al. demonstrated that insertion of a mutant *rpoB* gene into a MSSA strain led to conversion to MRSA [24]. Thus, mutations in both genes may contribute to the MODSA phenotype observed in this strain.

## Conclusion

In this study, we describe 2 different strains of MODSA mediated by two separate mechanisms. Subpopulations of the same strain of *S. aureus* in the same host but with different phenotypic and genotypic expressions, as opposed to subpopulations of differing strains, are not commonly seen. It is important to recognize that there are multiple mechanisms leading to MRSA that might be missed when testing only for the presence of MecA.

## Data Availability

Data have been deposited in Genbank.

## Acknowledgements

This work was funded by NIH/NIAID grant no. R01AI119145.

## References

1. Ba X, Kalmar L, Hadjirin NF, Kerschner H, Apfalter P, Morgan FJ, et al. Truncation of GdpP mediates β-lactam resistance in clinical isolates of Staphylococcus aureus. J Antimicrob Chemother. 2019;74:1182–91.

2. Argudín MA, Roisin S, Nienhaus L, Dodémont M, de Mendonça R, Nonhoff C, et al. Genetic Diversity among Staphylococcus aureus Isolates Showing Oxacillin and/or Cefoxitin Resistance Not Linked to the Presence of Genes. Antimicrob Agents Chemother [Internet]. 2018;62. Available from: http://dx.doi.org/10.1128/AAC.00091-18

3. Tomasz A, Drugeon HB, de Lencastre HM, Jabes D, McDougall L, Bille J. New mechanism for methicillin resistance in Staphylococcus aureus: clinical isolates that lack the PBP 2a gene and contain normal penicillin-binding proteins with modified penicillin-binding capacity. Antimicrob Agents Chemother. 1989;33:1869–74.

4. Jorgensen JH. Mechanisms of Methicillin Resistance in Staphylcoccus aureus and Methods for Laboratory Detection. Infect Control Hosp Epidemiol. Cambridge University Press; 1991;12:14–9.

5. Łeski TA, Tomasz A. Role of penicillin-binding protein 2 (PBP2) in the antibiotic susceptibility and cell wall cross-linking of Staphylococcus aureus: evidence for the cooperative functioning of PBP2, PBP4, and PBP2A. J Bacteriol. 2005;187:1815–24.

6. Banerjee R, Gretes M, Harlem C, Basuino L, Chambers HF. A mecA-Negative Strain of Methicillin-Resistant Staphylococcusaureus with High-Level β-Lactam Resistance Contains Mutations in Three Genes. Antimicrob Agents Chemother. American Society for Microbiology Journals; 2010;54:4900–2.

7. Ba X, Harrison EM, Edwards GF, Holden MTG, Larsen AR, Petersen A, et al. Novel mutations in penicillin-binding protein genes in clinical Staphylococcus aureus isolates that are methicillin resistant on susceptibility testing, but lack the mec gene. J Antimicrob Chemother. 2014;69:594–7.

8. Corrigan RM, Abbott JC, Burhenne H, Kaever V, Gründling A. c-di-AMP is a new second messenger in Staphylococcus aureus with a role in controlling cell size and envelope stress. PLoS Pathog. 2011;7:e1002217.

9. Griffiths JM, O’Neill AJ. Loss of function of the gdpP protein leads to joint β-lactam/glycopeptide tolerance in Staphylococcus aureus. Antimicrob Agents Chemother. 2012;56:579–81.

10. Göhring N, Fedtke I, Xia G, Jorge AM, Pinho MG, Bertsche U, et al. New role of the disulfide stress effector YjbH in β-lactam susceptibility of Staphylococcus aureus. Antimicrob Agents Chemother. 2011;55:5452–8.

11. CLSI. 2018. Performance standards for antimicrobial susceptibility testing, 28th ed., M100-S28E. CLSI,

12. Wayne PA Sullivan MJ, Altman DR, Chacko KI, Ciferri B, Webster E, Pak TR, et al. A Complete Genome Screening Program of Clinical Methicillin-Resistant Staphylococcus aureus Isolates Identifies the Origin and Progression of a Neonatal Intensive Care Unit Outbreak. J Clin Microbiol [Internet]. 2019;57. Available from: http://dx.doi.org/10.1128/JCM.01261-19

13. PacificBiosciences. PacificBiosciences/GenomicConsensus [Internet]. [cited 2020 Sep 16]. Available from: https://github.com/PacificBiosciences/GenomicConsensus

14. Li H. Minimap2: pairwise alignment for nucleotide sequences. Bioinformatics. 2018;34:3094–100.

15. Li H. Aligning sequence reads, clone sequences and assembly contigs with BWA-MEM [Internet]. arXiv [q-bio.GN]. 2013. Available from: http://arxiv.org/abs/1303.3997

16. Walker BJ, Abeel T, Shea T, Priest M, Abouelliel A, Sakthikumar S, et al. Pilon: an integrated tool for comprehensive microbial variant detection and genome assembly improvement. PLoS One. 2014;9:e112963.

17. Seemann T. Prokka: rapid prokaryotic genome annotation. Bioinformatics. 2014;30:2068–9.

18. Nicol JW, Helt GA, Blanchard SG Jr, Raja A, Loraine AE. The Integrated Genome Browser: free software for distribution and exploration of genome-scale datasets. Bioinformatics. 2009;25:2730–1.

19. Khelik K, Lagesen K, Sandve GK, Rognes T, Nederbragt AJ. NucDiff: in-depth characterization and annotation of differences between two sets of DNA sequences. BMC Bioinformatics. 2017;18:338.

20. Wang K, Li M, Hakonarson H. ANNOVAR: functional annotation of genetic variants from high-throughput sequencing data. Nucleic Acids Res. 2010;38:e164.

21. Camacho C, Coulouris G, Avagyan V, Ma N, Papadopoulos J, Bealer K, et al. BLAST+: architecture and applications. BMC Bioinformatics. 2009;10:421.

22. Pinho MG, de Lencastre H, Tomasz A. Transcriptional analysis of the Staphylococcus aureus penicillin binding protein 2 gene. J Bacteriol. 1998;180:6077–81.

23. Aiba Y, Katayama Y, Hishinuma T, Murakami-Kuroda H, Cui L, Hiramatsu K. Mutation of RNA polymerase β-subunit gene promotes heterogeneous-to-homogeneous conversion of β-lactam resistance in methicillin-resistant Staphylococcus aureus. Antimicrob Agents Chemother. 2013;57:4861–71.

24. Panchal VV, Griffiths C, Mosaei H, Bilyk B, Sutton JAF, Carnell OT, et al. Evolving MRSA: High-level β-lactam resistance in Staphylococcus aureus is associated with RNA Polymerase alterations and fine tuning of gene expression. PLoS Pathog. 2020;16:e1008672.

